# Framework for precision medicine in focal segmental glomerulosclerosis: Translation of sparsentan-responsive genes in a rat model to kidney disease associated proteins in biofluids

**DOI:** 10.1101/2025.02.26.25322958

**Authors:** Sean Eddy, Viji Nair, John Hartman, Damian Fermin, Felix Eichinger, Bradley Godfrey, Wenjun Ju, Jeffrey B Hodgin, Laura H. Mariani, Patricia W. Bedard, Celia P. Jenkinson, Bruce Hendry, Radko Komers, Jula Inrig, Matthias Kretzler, Nephrotic Syndrome Study Network (NEPTUNE)

## Abstract

Sparsentan, a dual endothelin receptor A inhibitor and angiotensin blocker, reduced proteinuria in patients with focal segmental glomerulosclerosis (FSGS) in Phase II and Phase III studies. However, the estimated glomerular filtration rate (eGFR) endpoint was not achieved, partially attributed to disease heterogeneity among participants. Sparsentan reversed the molecular fingerprint in kidneys of an adriamycin-challenged rat model of chronic kidney disease, consistent with the phenotypic data. Transcriptomic profiles from this model were used to identify differentially expressed genes, 388 of which had reversed directionality, and networks responsive to sparsentan. These included disease networks relevant to FSGS, including suppression of cytokine signaling, immune and inflammatory pathways as well as inhibition of networks downstream of endothelin and angiotensin activation. Human orthologs of genes upregulated and reversed by sparsentan in the rat model were elevated in glomerular (p<0.001) and tubulointerstitial (p<0.0001) profiles of participants with FSGS in the Nephrotic Syndrome Study Network (NEPTUNE) compared to healthy living donors, pointing to a likely anti-inflammatory action of sparsentan on kidneys. The gene signature in both compartments was negatively correlated with eGFR (p<0.005) and positively correlated with UPCR (p<0.005) and the response profile was elevated in a molecular subgroup of patients with greater disease severity. Several urine proteins were associated with the sparsentan response score highlighting opportunities for the development of non-invasive surrogates of sparsentan response to enable a precision medicine approach for treatment with dual endothelin angiotensin receptor antagonists.

**Translational Statement:** Cross-species mapping of sparsentan response in rat and human studies identified similar networks which were elevated in a subset of people with more severe focal segmental glomerulosclerosis (FSGS) providing the basis for implementing precision medicine in for sparsentan treatment.

## Introduction

Focal segmental glomerulosclerosis (FSGS) is a heterogeneous syndromic pathology characterized by glomerular lesions that lead to both segmental scarring, and podocyte injury. It is the underlying cause of nephrotic syndrome in nearly 20% of pediatric cases, and 40% of adult cases^1^. Although a rare disease, in the United States, it is the most common primary glomerular disorder that leads to end-stage renal disease^2^. The economic burden of FSGS is substantial with >$68,000 per year per patient with kidney failure attributed to FSGS^3^.

To date, no pharmacological therapy has been approved specifically for FSGS. The Kidney Disease: Improving Global Outcomes (KDIGO) guidelines^4^ recommend a tiered approach for treating FSGS. involving renin-angiotensin system inhibitors (RASi) either angiotensin-converting enzyme (ACE) inhibitors or angiotensin receptor blockers (ARBs) to control proteinuria ^5^, and often high dose steroids and other immunosuppressive therapies (IST) in an attempt to achieve disease remission. Novel therapies targeting disease mechanisms active in patients with FSGS, with fewer side-effects than available IST, that can prevent or delay disease progression are much needed.

Angiotensin II (Ang II) and endothelin-1 (ET1) are vasoactive peptide mediators with multiple physiological and pathophysiological actions in the kidney. Simultaneous inhibition of both peptides has been shown to be more nephroprotective than treatments with inhibitors of each peptide alone in various models of kidney diseases including FSGS as well as in clinical studies of patients with diabetic and non-diabetic nephropathies^6, 7^.

Sparsentan is a single molecule dual antagonist of endothelin type A (ET_A_) and Ang II type 1 (AT_1_) receptors^7^ Similar to a treatment with two monoblockers, sparsentan reduced proteinuria, expression of glomerulosclerosis markers, renal fibrosis, and inflammation in several models of kidney disease^7^. In clinical studies, sparsentan reduced proteinuria in patients with FSGS in an 8 week Phase II randomized, double-blind, active control study (DUET)^8^, and in a 108 week Phase III study (DUPLEX) in comparison to irbesartan^9^, although the antiproteinuric effect did not translate into a statistically significant, clinically meaningful differences in eGFR slope were observed over 2-years . ^9^.

To further characterize the potential for sparsentan to treat patients with FSGS, we undertook a cross-species transcriptomic study to identify a cross-species conserved pharmacodynamic responses in kidneys. Using a rat adriamycin (ADR) model of CKD, which displays a FSGS-like phenotype^10–12^ , Animals were treated with sparsentan or vehicle control. Sparsentan-responsive transcriptional profiles in ADR rats were mapped onto transcriptional profiles from micro- dissected glomeruli and tubulointerstitia from kidney biopsies available in the Nephrotic Syndrome Study Network (NEPTUNE) ^13–15^, from patients with FSGS.

## Methods

### Animals

Male Sprague Dawley rats (11-13 weeks old) were acclimated 7 days prior to ADR administration (5 mg/kg, single IV injection) and followed for another 7 days allowing the development of proteinuria. Eight days after ADR challenge, the animals were randomized to receive vehicle (ADR-Veh, n=10) or sparsentan 6 mg/day (ADR-Spa6, n=10), 18 mg/day (ADR-Spa18, n=10), or 60 mg/day (ADR-Spa60, n=9) by daily oral gavage for 35 days. Urine protein creatinine ratios (UPCR) were measured from 24 hr urine samples taken prior to ADR disease induction, at randomization, and on days 14 and day 33 after initiation of sparsentan dosing. Sham/Vehicle (Veh, n=5) animals received no ADR challenge. Tissues were collected 36 days after dosing began.

### Urine protein creatine ratio and glomerulosclerosis severity scores

24 hr urine samples were analyzed for total protein using the Roche Total Urine Protein assay (Indianapolis, U.S.). Whole kidneys were fixed in 10% neutral buffered formalin and cut in 2-3 mm coronal slices, returned to formalin and fixed overnight at 4°C before embedding in paraffin using standard histological processes. Cut sections were stained by periodic acid-Schiff hematoxylin (PASH) , histopathological analysis and images were taken using an Olympus BX51 research microscope equipped with a DP-71 digital camera. Glomerular lesions were evaluated utilizing a semi-quantitative assessment using the following scale. Normal = 0; within normal limits (no pathological change); Mild = 1+; up to ½ glomerular structure is affected; Moderate = 2+; up to ¾ glomerular structure is affected; Severe = 3+; global involvement of all glomerular structure.

### Human data

NEPTUNE (NCT0120900) is a prospective study of children and adults with proteinuria currently recruited from 31 sites at the time of their first clinically indicated kidney biospy^15^.

Informed consent was obtained from individual patients or parents/guardians. The study was approved by the institutional review board at the University of Michigan. Clinical data (eGFR and UPCR) were extracted from the July 28, 2022 NETPUNE database.

### FFPE fixation of kidneys and RNA extraction

Rat kidney harvested and fixed in FFPE blocks. FFPE sections were microtome into 20µm rolls. Deparaffinization solution (Qiagen, Cat. #19093) was used to remove paraffin prior to proteinase K digestion according to manufacturer’s protocols. Total RNA was isolated using RNeasy FFPE kits (Qiagen, cat# 73504) according to manufacturer’s protocols. QC analysis was performed on an Agilent Bioanalyzer 2100.

### Transcriptomic analysis of Rat Kidney RNA

RNA sequencing (RNA-seq) library preparation and sequencing was performed at the University of Michigan Sequencing Core using a Novaseq 4000 (Illumina). RNA-seq reads were aligned to Rattus norvegicus genome build Rnor 6.0.88. QC was performed to identify any technical outliers and counts were aggregated at the EnsemblID gene level. One ADR animal treated with 60 mg/kg did not pass technical QC. RNA-seq profiles were analyzed using DESeq2^16^ after filtering out low count genes (<1 CPM in at least 2 samples) in a locally installed version of DEApp^17^. The data/analyses presented in the current publication will be deposited in and are available from the dbGaP database and matrix gene counts will be available in GEO.^18, 19^

### Sparsentan score in kidney biopsies

Glomerular (n=93) and tubulointerstitial (n=114) compartment-enriched genome-wide mRNA expression profiles were generated from kidney biopsies of NEPTUNE^13^ participants with biopsy-proven FSGS and from comparator healthy living transplant donors and are deposited into GEO as part of NEPTUNE transcriptomic datasets GSE254957 and GSE219185.

Human-rat ortholog mapping was performed using ENSEBML Biomart (v104).^20^ Human orthologs of genes responsive to sparsentan in rats were evaluated in patients with FSGS from the NEPTUNE^13, 15^ (Supplementary Table 1). Genes that were up-regulated in the model and then down-regulated by sparsentan at the 60mg/kg dose were used to generate a sparsentan score in human kidney biopsy voom-transformed gene expression profiles in healthy living donors and in patients with FSGS. Briefly, each gene profile was converted to a Z-score, and the average Z-score of all these genes was computed as the sparsentan score, as previously described for TNF^13^ in the transcriptional datasets noted above.

### Statistical analysis

#### ADR rat model data

A two-way ANOVA mixed model was used to determine significant differences in UPCR between vehicle treated animals and ADR-challenged animals and between ADR-challenged animals and animals sparsentan intervention. A one-way ANOVA test was used to determine significant differences in severity of glomerular injury.

#### Sparsentan scores in participants with FSGS

The sparsentan score was correlated with centrally measured UPCR +1 (log2 transformed) and estimated glomerular filtration rate (eGFR) closest to biopsy^14^using Pearson’s correlation. A Welch’s t-test was used to determine significance between sparsentan score profiles from healthy living transplant donors and patients with FSGS. Statistical tests were applied in GraphPad Prism v10.2.2.

### Somascan profiling

Urinary proteins were quantified using SomaScan’s 7k (SomaLogic) aptamer-based profiling assay^21^ in urine samples from NEPTUNE patients closest to kidney biopsy. Relative fluorescent units representing each aptamer were generated and extracted in R using the SomaDataIO package (https://somalogic.github.io/SomaDataIO/) and log2 transformed. For QC, probes were identified with signal above background using the Wilcoxon Rank Sum test and the estimated limit of detection^22^. The number of analytes passing both QC parameters in urine was 1,183. Lastly, values were normalized to urine creatinine.

## Results

### Sparsentan reduces ADR driven proteinuria and glomerulosclerosis in a dose-responsive manner

Overview of the study design is shown in Figure 1. Sham/vehicle treated (Veh) animals had no measurable UPCR. ADR challenged, vehicle treated (ADR-Veh) rats developed severe proteinuria compared to Veh animals after day 14 which continued through to the end of study (p<0.0005 and p<0.0001 respectively, Figure 2A), This was attenuated by treatment with sparsentan, resulting in a significant reduction in proteinuria in the highest dose group (p<0.05 ADR-Spa60 vs. ADR-Veh, Figure 2 A)). Similarly, Veh treated animals had no evidence of glomerulosclerosis. In contrast, ADR-Veh rats developed marked glomerulosclerosis, which was dose-dependently reduced in ADR-Spa18 (p<0.05) and ADR-Spa60 rats (p<0.0005)(Figure 2B).

**Figure 1.**
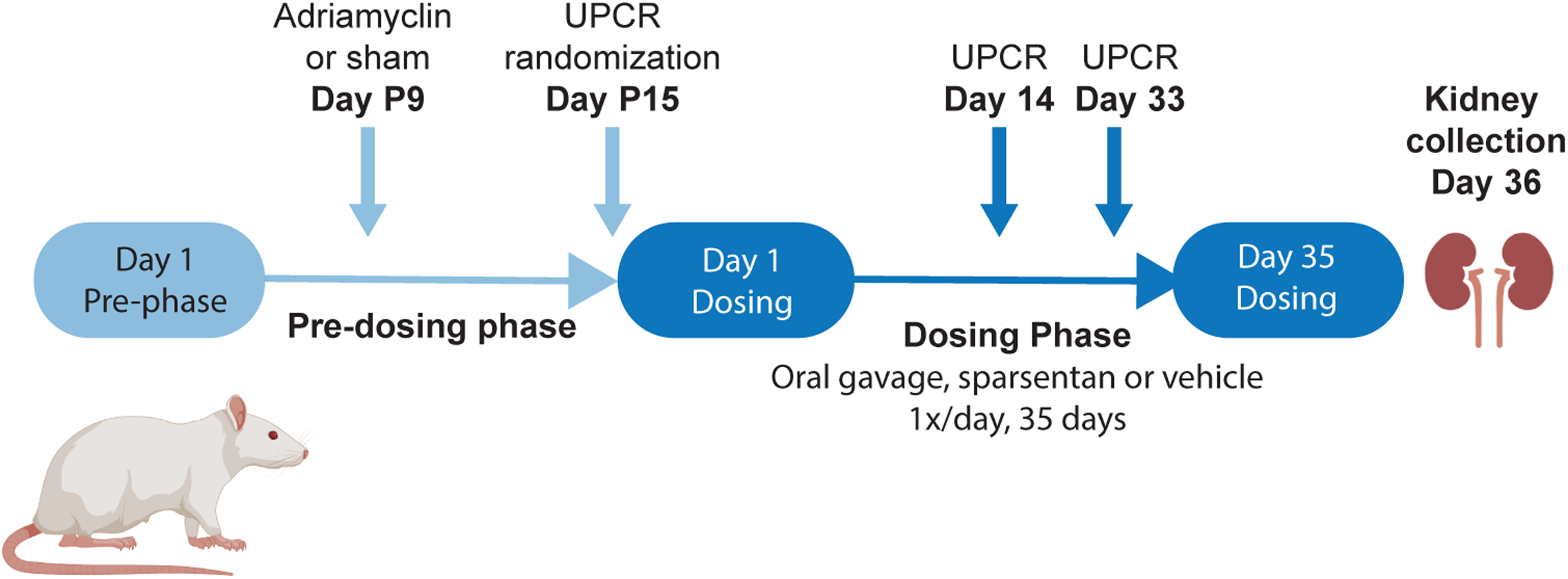
Study design and dosing model to assess the ability of sparsentan to attenuate an FSGS-like disease phenotype driven by adriamycin in rats. Male Sprague Dawley rats, 11-13 weeks old at start were treated with 5mg/kg Adriamycin (ADR) by single IV injection or vehicle (sham) on day P9 followed by sparsentan treatment after 6 days. UPCR was measured on days 14 and 33 of sparsentan treatment. Kidneys were harvested one day after final treatment.

**Figure 2.**
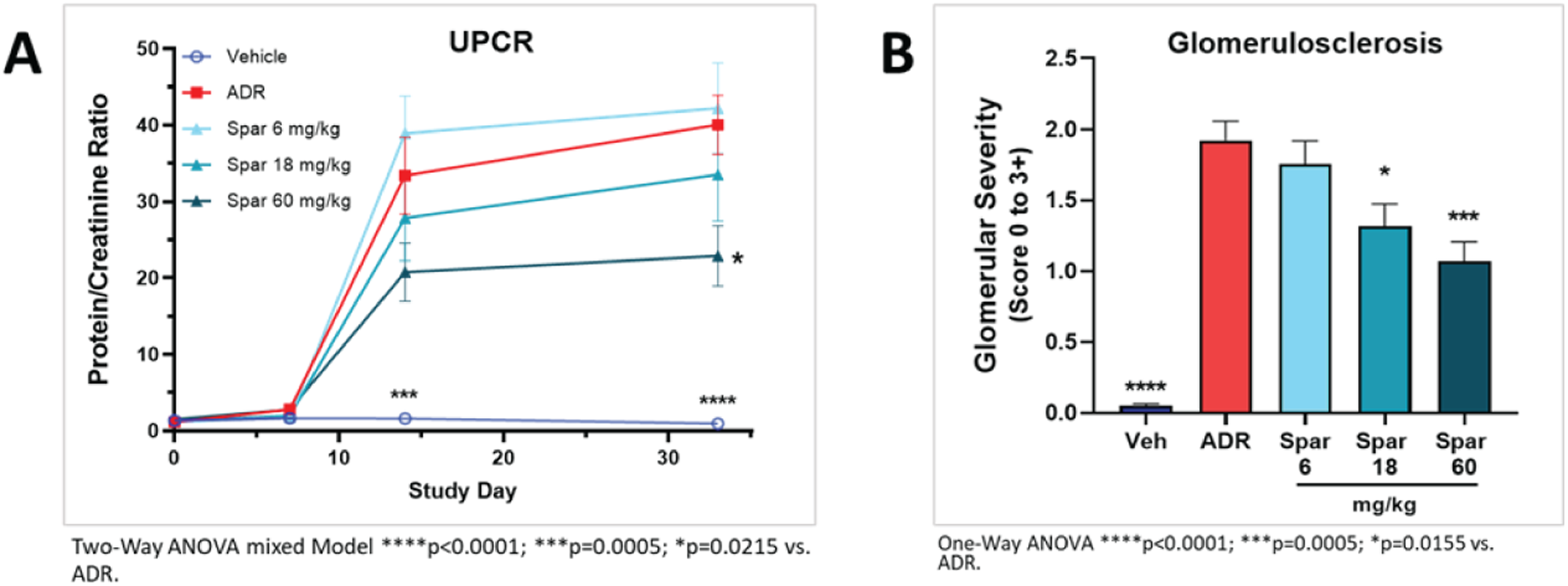
Sparsentan treatment effects in ADR rats. **a.** Dose response of urine protein to creatinine ratio (UPCR) across the study timeline and **b.** glomerulosclerosis quantified at the end of the study in the ADR rat model.

### Impact of ADR and sparsentan on transcriptional profiles in rat kidneys

To determine if the phenotypic effects of reduced UPCR and lower glomerulosclerosis after sparsentan intervention affected transcriptional networks, we extracted RNA from paraffin-embedded whole kidney tissue and generated RNA count profiles from Veh, ADR-Veh, ADR- Spa6, ADR-Spa18, and ADR-Spa60 experimental rat groups. After filtering out low count genes, 17,490 ENSEMBL genes remained for differential expression analysis.

First, we established a baseline understanding of differentially expressed gene (DEG) from disease induction by ADR in rat kidneys by comparing ADR-Veh to Veh animals. In this comparison, 4280 genes were differentially expressed (2271 up-regulated and 2009 down- regulated, p-adj<0.05) (Table 1 and volcano plot, Supplemental Figure 1a). Among the top significant up-regulated DEGs (by fold change (FC)) in kidneys from ADR-Veh animals compared to Veh animals were *Il19* (Log_2_ FC=8), *Havcr1* (Log_2_ FC=5.8), and *Il24* Log_2_ FC=5.8). Among 2271 up-regulated genes, enrichment analysis using Enrichr ^23–25^ and the Reactome database (version 2022)^26–28^ yielded 2250 mappable genes, which were consistent with highly enriched activated immune and cytokine signaling and extra cellular matrix-related pathways (Supplemental Figure 1C). Among the 2009 down-regulated genes, 2008 were mapped for enrichment analysis, which overrepresentation of genes in multiple metabolic pathways (Supplemental Figure 1D).

**Table 1.** The number of significantly differentially expressed genes (p-adj<0.05) in each differential expression analysis comparison using DESeq2.

| Comparison | Number of genes |  |
| --- | --- | --- |
|  | up-regulated | down-regulated |
| ADR-Veh vs. Veh | 2271 | 2009 |
| ADR-Spa60 vs. ADR-Veh | 290 | 292 |
| ADR-Spa18 vs. ADR-Veh | 4 | 0 |
| ADR-Spa6 vs. ADR-Veh | 0 | 39 |

To establish the impact of sparsentan on transcriptome-wide expression profiles resulting from disease induction, differential expression analysis was performed on RNA-seq profiles from kidneys of each of the treated animal groups (ADR-Spa6, ADR-Spa18, and ADR-Spa60) compared to RNA-seq profiles of kidneys from ADR-Veh animals. In the ADR-Spa6 vs. ADR and ADR-Spa18 vs. ADR comparisons, fewer than 50 differentially expressed genes were detected (Table 1). The limited impact of sparsentan on the transcriptomes in the kidneys of ADR-Spa6 and ADR-Spa18 compared to ADR-Veh animals was consistent with UPCR findings from animals in these treatment groups. Lastly, ADR-Spa60 compared to ADR-Veh resulted in 582 differentially expressed genes in the kidney (290 up-regulated with sparsentan, 292 down- regulated, p-adj<0.05) (Table 1 and volcano plot, Supplemental Figure 1b). The highest upregulated gene was *Ren* (Log_2_ FC=9.2), consistent with blockade of AT_1_ receptor activity ^29–32^

Among the top up-regulated genes in the model (*Il24*, *Havcr1,* and *Il19*), *Il24* and *Havcr1* were both significantly repressed (p-adj<0.05) in ADR-Spa60 animals compared to ADR-Veh animals (Log_2_ FC=-3.0 and Log2 FC=-1.9, respectively), while *Il19* had a trend of down-regulation (Log_2_ FC=-2.2, p-adj=0.067). By pathway enrichment, the 292 genes repressed by sparsentan were consistent with suppression of cytokine signaling, immune and inflammatory pathways (Supplemental Figure 1F).

### Impact of sparsentan on model-induced gene expression profiles

Although we only observed a limited number of significant DEGs in the lower dose comparisons (ADR-Spa6 vs. ADR-Veh and ADR-Spa18 vs. ADR-Veh), we wanted to better assess whether there was an overall dose-dependent response associated with sparsentan. To achieve this, we used differential expression results from all 17,490 genes in each differential expression comparison (independent of significance) and performed a linear regression analysis and plotted results between log_2_ fold changes from ADR-Veh compared Veh against ADR-Spa6 (left plot, Supplemental Figure 2, r=-0.09), ADR-Spa18 (middle plot, Supplemental Figure 2, r=-0.47) and ADR-Spa60 (right plot, Supplemental Figure 2, r=-0.57) . As demonstrated by the correlation coefficients, and consistent with selected genes reported above, sparsentan reverses the overall transcriptional landscape from ADR-challenge, although this reversion is incomplete (see Log_2_ fold changes in Supplemental Figures 1A, 1B, and Supplemental Figure 2).

Looking specifically at significant DEGs (Table 1), 67% (389/582) of genes significantly differentially expressed in the ADR-Spa60 vs. ADR-Veh comparison were also differentially expressed in the ADR-Veh vs. Veh comparison (p-adj<0.05, see Venn diagram, Figure 3A). All but one gene (*Ccng1*) of the 389 shared genes reversed directionality of gene expression compared to the ADR-Veh vs. Veh comparison (Figure 3B); genes with positive fold changes in the ADR-Veh vs. Veh comparison had negative fold changes in the ADR-Spa60 vs. ADR-Veh comparison and vice versa. Of the 388 genes with reversed directionality, 212 genes were up- regulated in the ADR-Veh vs. Veh comparison and down-regulated in the ADR-Spa60 vs. ADR- Veh comparison, while 176 were down-regulated in the ADR-Veh vs. Veh comparison and up- regulated in the ADR-Spa60 vs. ADR-Veh comparison. As noted, *Ccng1*, the lone exception, was up-regulated ADR-Veh vs. Veh comparison (Log2 FC=0.39, p-adj<0.05) and also up- regulated in the ARD-60 vs ADR-Veh comparison (Log2 FC=0.25, p-adj<0.05). Taken together, these data support the ability of sparsentan to reverse the molecular fingerprint of ADR- challenge in rat kidneys, particularly at higher doses, consistent with the phenotypic data for animals in this study.

**Figure 3.**
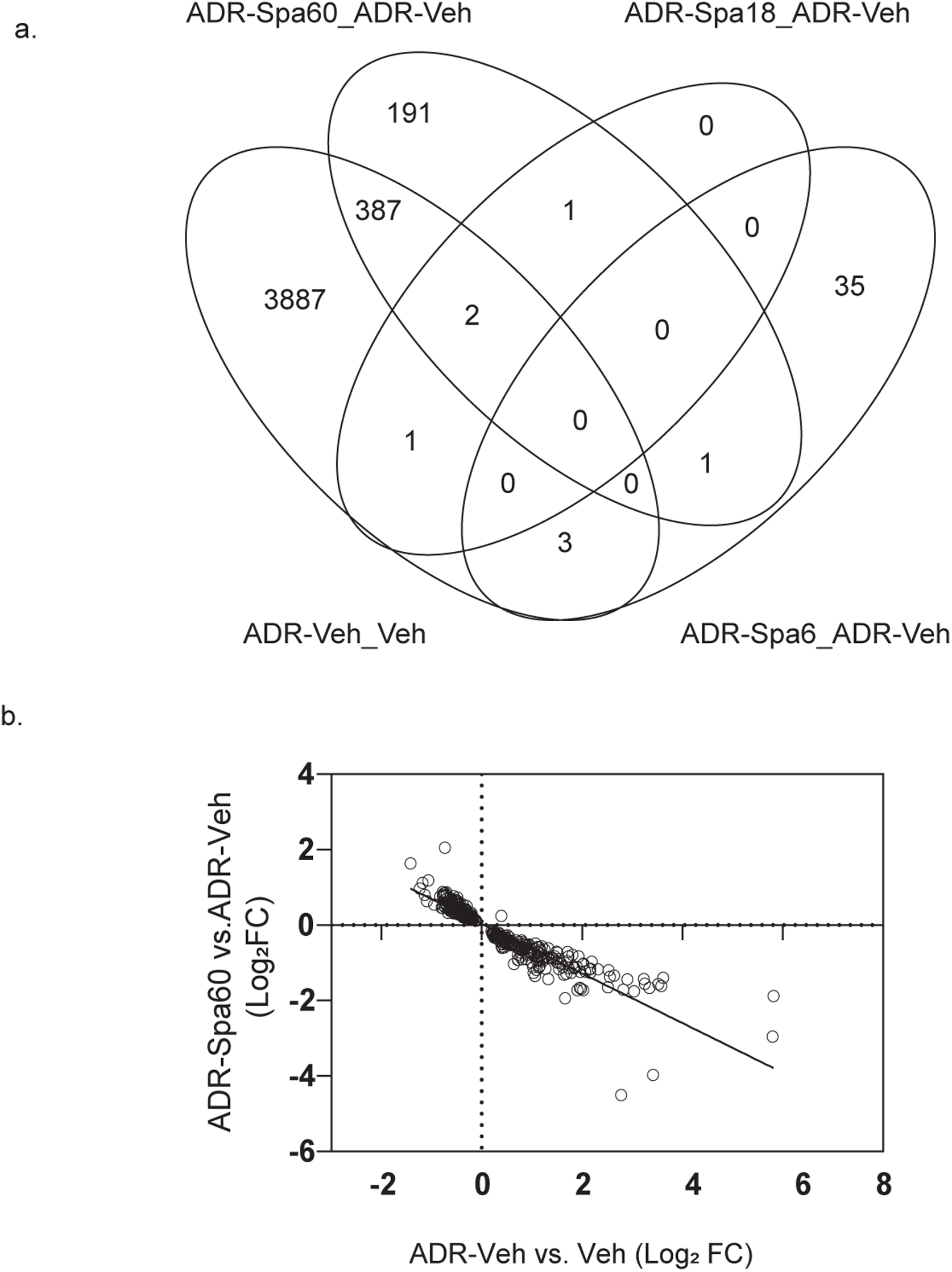
Transcriptional changes with sparsentan treatment in ADR rats. **a.** A majority of significant differentially expressed genes (DEGs, p-adj<0.05) in the ADR-Spa60 vs ADR-Veh comparison (389/582) were found in the ADR v Sham comparison. **b.** Directionality of the 389 genes common between the two comparisons were reversed, consistent with attenuation of the disease signal by sparsentan.

### Upstream regulator analysis reveals reversal of disease mechanisms with sparsentan

Differentially expressed genes for the ADR-Veh compared Veh comparison and the ADR-Spa60 compared ADR-Veh were analyzed for predicted upstream regulator activity (IPA). Upstream regulators are potential causal explanations for the observed differential gene expression values in each analysis^33^. Only genes with a |fold change| >1.5 and p-adj<0.05 that mapped into the upstream regulator database were considered. ^29–31^. Top predicted upstream regulators identified in the ADR-challenged model included a number of human FSGS-relevant mechanisms included inflammatory, fibrotic, and immune mediators, TNF^33, 34^, TGFB^34^, IL1B^34^, IFNG^34^ (Table 2).

**Table 2.** IPA upstream regulator networks identified from DEG profiles.

| Upstream regulator | ADR vs. sham | Sparsentan High vs. ADR |
| --- | --- | --- |
|  | Z-score | Z-score |
| TNF | 10.5 | -4.4 |
| TGFB1 | 8.3 | -2.7 |
| IFNG | 7.6 | -2.5 |
| IL1B | 6.9 | -2.9 |
| CSF2 | 8.3 | -3.6 |

Upstream regulator analysis also predicted activation of both AGT (angiotensinogen) and EDN1 (endothelin-1) in the ADR-Veh vs. Veh comparison (Activation Z-scores of 8.2 and 4.0, respectively), while activation Z-scores consistent with a reduction in predicted activity were found in the ADR-Spa60 vs. ADR comparison for both mechanisms (-1.4 and -1.6, respectively), consistent with the dual antagonist mechanism of action of sparsentan.

### Sparsentan responsive genes were associated with human FSGS and clinical measures of disease

With the animal models highlighting mechanisms relevant to human disease and to the mechanism of action of sparsentan, we next sought to understand whether the transcriptional networks themselves were directly implicated in human disease. Because the majority of the 388 genes reversing direction with sparsentan treatment were up-regulated by ADR-challenge but down-regulated by sparsentan (lower left quadrant in Figure 3B), we focused our attention on these 212 genes. Using ENSEMBL Biomart (v104) ortholog mapping^20^, the 201 of the 212 rat genes had human orthologs and mapped to 197 human Ensembl genes (due to instances of multiple rat genes mapping to a single human gene, Supplemental Table 1). We computed a network activity score from human orthologs expressed in glomerular and tubulointerstitial transcriptomes for each patient, using methods previously reported for TNF activation^13^, to obtain the sparsentan score. The sparsentan score therefore represents a network of human gene orthologs of genes induced in the kidney of the ADR rat model and repressed by sparsentan intervention. A high sparsentan score is indicative of elevated gene network in humans, that was also reversible by sparsentan in a model system, while a low score indicates lower or lack of activation of this network. We focused on comparing transcriptional profiles of biopsies from healthy living transplant donors and patients with FSGS (FSGS demographics presented in Table 3) and hypothesized that the sparsentan score would be elevated in patients with FSGS. The sparsentan score was indeed elevated in both glomerular (glom) and tubulointerstitial (TI) transcriptional profiles from biopsies of patients with FSGS compared to healthy living transplant donors (Figure 4A, p<0.001 in glom, Figure 4B, p<0.0001 in TI), was negatively correlated with eGFR (Figure 4C, R=-0.31, p<0.005 in glom and Figure 4D, R=-0.42, p<0.0001 in TI), and displayed a modest positive correlation with log_2_ UPCR +1 (Figure 4E, R=0.33, p<0.005 in glom and Figure 4F, R=0.29, p<0.005 in TI). Lastly, the sparsentan score was elevated in TI from patients with a molecularly driven inflammatory TNF cluster (Cluster 3, Figure 4G, p<0.0001 vs Cluster 1 and Cluster 2 or Cluster 1 and 2 combined) previously characterized as associating with poor outcome and a high intrarenal inflammatory state ^13^. Thus, the molecular profile reversed by sparsentan in a model system was consistent with an active and inflammatory disease profile in patients with FSGS pointing to a likely anti-inflammatory action of sparsentan in kidneys.

**Figure 4.**
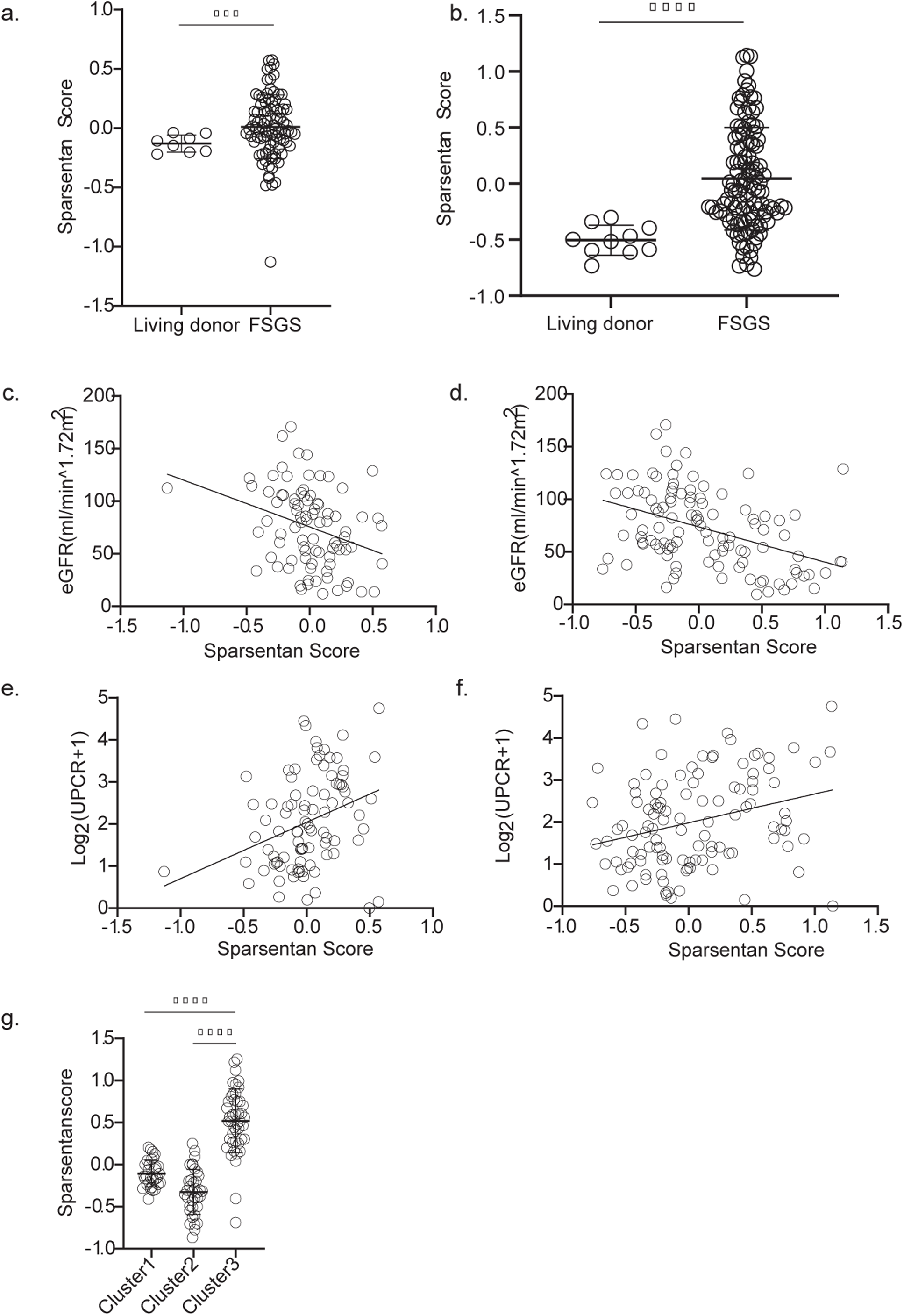
Sparsentan responsive genes in human data. Human orthologs of genes suppressed by sparsentan in the ADR-Spa60 vs ADR-Veh comparison (lower right quadrant, Figure 3b) were Z-transformed and the average of all genes was used to compute an intrarenal sparsentan response score from glomerular (glom) and tubulointerstitial (TI) transcriptomes. The sparsentan response score was elevated in patients from NEPTUNE with FSGS compared to healthy living transplant donors in the **a.** glom and **b.** TI. The score was negatively correlated with eGFR at time of biopsy in the **c.** glom and **d.** TI, and positively correlated with UPCR at time of biopsy in the **e.** glom and **f.** TI. **g.** The sparsentan score was also elevated in the TI from patients with FSGS that were part of a previously characterized TNF cluster (cluster 3).

**Table 3.**
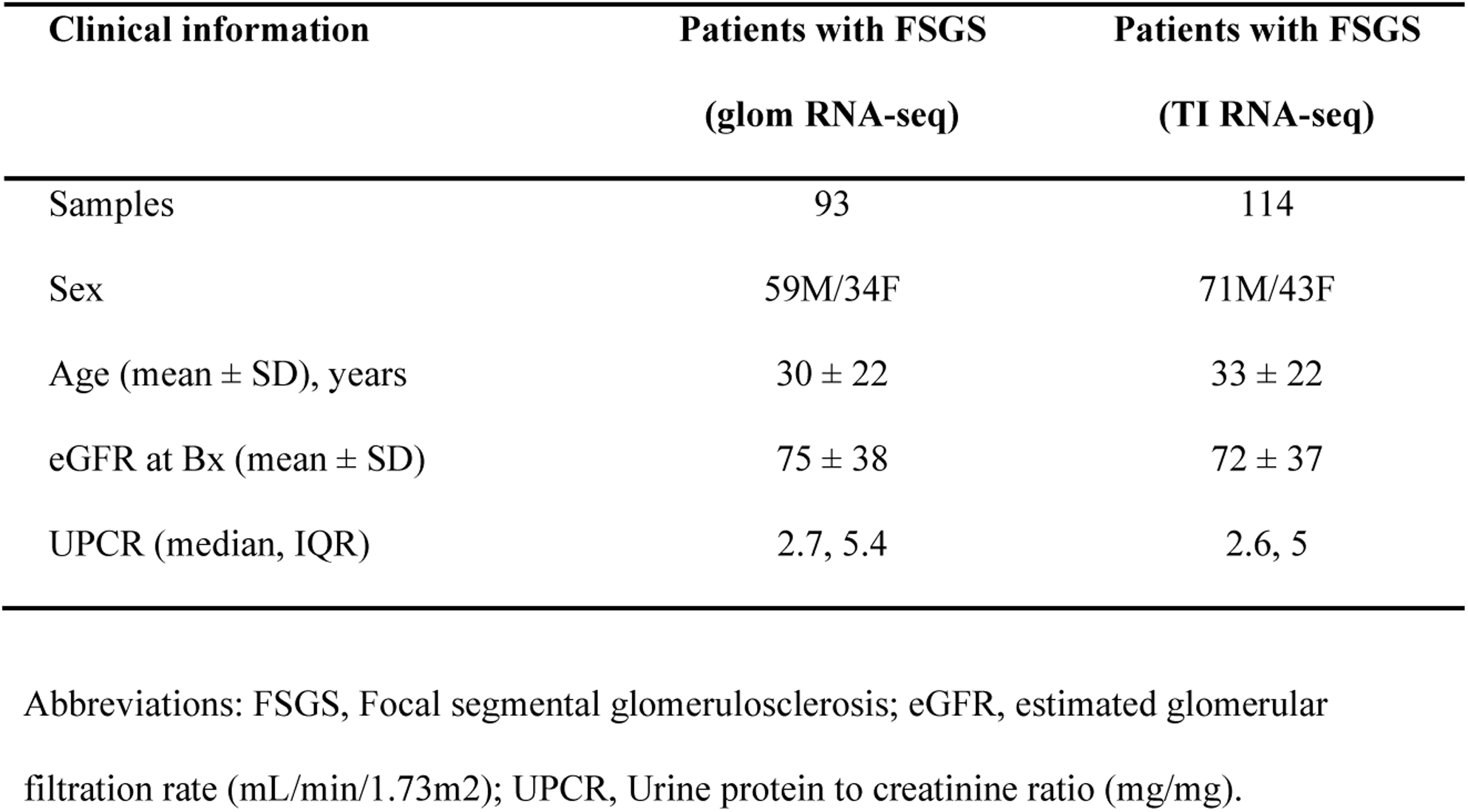
Time of biopsy clinical information for patients diagnosed with FSGS.

### Urinary protein profiles were strongly associated with glomerular and tubulointerstitial sparsentan scores in patients with FSGS

To determine the possibility of a noninvasive measure of sparsentan score activation, sparsentan scores from the glomerular and tubulointerstitial transcriptomes of patients with FSGS were correlated with their urine Somascan profiles (1,813 Aptamers) normalized to creatinine. In 35 paired glomerular-urine samples from patients with FSGS, a majority of urine aptamers (1,437) strongly correlated with glomerular sparsentan scores (R>0.4, FDR<0.05, Table 4). The Somascan platform included 27 aptamers corresponding to genes used to compute the sparsentan score. Of these, 23 were among the 1,437 strongly correlated aptamers. In 42 paired tubulointerstitial-urine samples from patients with FSGS, a majority of urine aptamers were also strongly correlated with the tubulointerstitial sparsentan score (R>0.4, FDR<0.05, Table 4).

**Table 4.** Correlation of sparsentan scores with urine Somascan profiles.

| **Sparsentan score** | **Samples with** | **Probes** | **Sparsentan score probes** |
| --- | --- | --- | --- |
|  | **urine profiles** | **(FDR<0.05, R>0.4)** | **(FDR<0.05, R>0.4)** |
| Glomeruli | 35 | 1437 | 23 |
| Tubulointerstitium | 42 | 1408 | 21 |

These included 1,313 that were in common with glomerular correlated urine probes (Figure 5A). As the aptamers correlated with the sparsentan scores from glom and TI were largely overlapping, we took the average correlation coefficient of probes. The top 2 correlated probes corresponded were complement component C7 and Notch receptor 3 and among genes that contributed to the sparsentan signature, probes *FGB* (fibrinogen beta chain) and *UNC5B* (unc-5 netrin receptor B). Correlation plots for these probes against the sparsentan scores (TI) are shown in Figures 5B-D. Thus, these probes could be used to monitor or evaluate the sparsentan score noninvasively.

**Figure 5.**
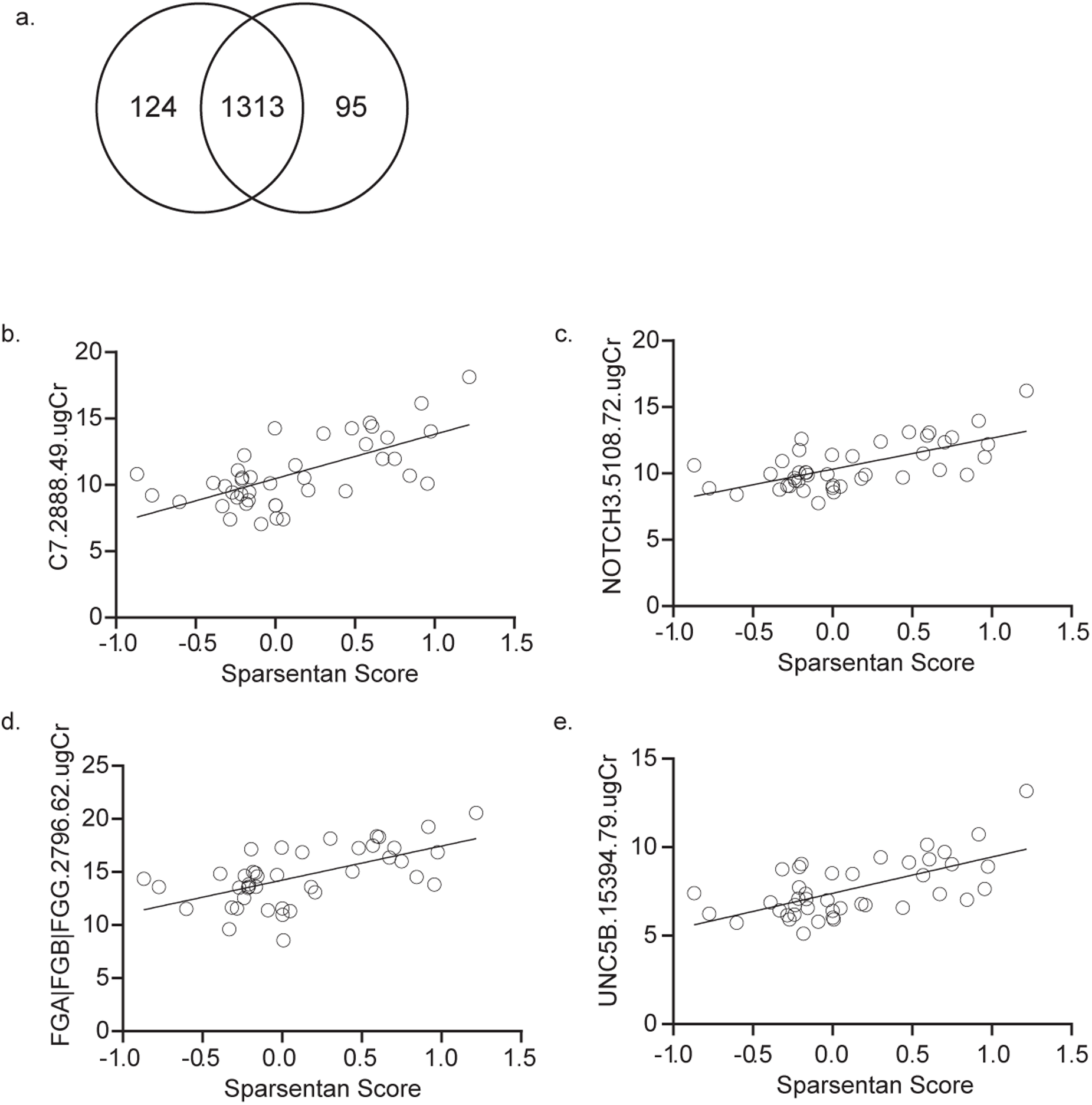
The intrarenal sparsentan scores were correlated with a large number of urine protein probes in patients with FSGS. **a.** Urine probes correlated (R>0.4, FDR<0.05) with glomerular (glom) and tubulointerstitial (TI) sparsentan scores were largely overlapping. The top urine probes correlated with both glom and TI sparsentan scores were plotted against the TI sparsentan score; C7 **b.** (r=-0.65, p-adj=0.001) and NOTCH3 **c.** (r=-0.66, p-adj=0.001). The top urine probes that were part of the sparsentan score and correlated with both glom and TI sparsentan scores were plotted against TI sparsentan score, **d.** FGB (r=-0.60, p-adj=0.001) and **e.** UNC5B (r=-0.63, p-adj=0.001).

## Discussion

FSGS is a rare disease, with an estimated incidence of 2.7 new cases per 100,000 people per year in the United States^35^. Despite being classified as a rare disease, it is one of the most common causes of end-stage renal disease^2^. Because FSGS is diagnosed based on histopathological descriptions of ultrastructural glomerular injury patterns in the kidney, despite a broad spectrum of processes and disease drivers, biopsy driven diagnosis provides an incomplete view for clinicians to make accurate treatment decisions. As such, histologic diagnoses alone cannot be expected to identify patients that will respond to treatments with defined specific mechanism of action in a uniform manner. To begin addressing this, we identified a transcriptional profile in an FSGS model that was reversed by sparsentan and performed a cross-species comparison from transcriptional profiles from human kidney biopsies to determine if similar networks were activated in patients with FSGS.

ADR-challenged rats are an established model of FSGS which develops podocyte injury as well as structural and functional changes reminiscent of FSGS in humans^36^. In this study, ADR- challenged rats displayed dose-dependent beneficial effects of sparsentan on the development of glomerulosclerosis and proteinuria, with statistically significant differences in ADR-60 rats compared with ADR-Veh, which was also reflected in DEG analyses with the highest number of significant DEGs between these groups. Therefore, further analyses were performed using the ADR-60 vs, ADR-Veh samples. To have a better mechanistic understanding of sparsentan’s actions in human FSGS, we performed a cross-species analysis using transcriptomic profiles generated from NEPTUNE.

NEPTUNE is designed as a multicenter collaborative study to establish the translational infrastructure necessary to carry out molecularly defined studies in nephrotic syndrome, including patients with biopsy-proven FSGS^15, 37^. At a high level, cross-species analysis revealed an elevated signaling profile in patients with FSGS, consistent with an activity profile amenable to sparsentan intervention. Nevertheless, even within patients with FSGS, a tremendous degree of heterogeneiety was observed, with some patients comparable to healthy living transplant donors and others with activated signaling with regards to gene networks impacted by sparsentan. This may allow for stratification of patients with FSGS either through identification and refinement of biomarkers associated with sparsentan response, or through further refinement of sparsentan- responsive signatures by linking activity profiles to better defined etiologies of FSGS lesions, thus reducing the heterogeneity of studied populations.

This study revealed several expected and some less expected DEGs in rats that were targeted by sparsentan treatment. These DEGs included *Ren*, kidney injury markers such as *Havcr1* (encoding Kim-1), and *Il24*, but also included gene networks related to inflammation and fibrosis (e.g. TNF, IFNG, TGFB1, IL1B) indicating broad impacts of these networks by sparsentan. These results correspond to previous abundant evidence in models of kidney disease that demonstrated similar effects of dual AT1 and ETA receptor inhibition on these molecular markers of kidney injury in parallel with amelioration of renal structural and functional alterations [refs 6.7]. The gene network reversed by sparsentan was elevated in human FSGS, negatively correlated with cross-sectional eGFR, associated with elevated proteinuria and found prominently elevated in a molecular cluster of FSGS patients associated with inflammatory TNF-associated disease^13^, suggesting broader effects of sparsentan beyond direct endothelin and angiotensin antagonism.

Disease heterogeneity and low incidence of FSGS underscores the need to develop a precision medicine framework in FSGS^37^. An increasing number of new therapies is in clinical development for FSGS^38^. Identifying potential responder populations in this heterogeneous indication for given therapies will be crucial for both clinical trial success and acceptance of new therapies in the FSGS community. In this paper, we report the process to elucidate clinically relevant signaling in patients with FSGS using the dual AngII/ET-1 receptor inhibitor sparsentan in model systems. Moreover, this signaling network was elevated in a molecular subset of patients previously identified at a higher risk for progressive loss of kidney function^13^. These patients might be specifically indicated for sparsentan treatment. However, high number of DEGs reversed by sparsentan treatment as observed in the ADR model indicates a therapeutic potential of sparsentan in a broad FSGS population. This is supported by clinical experience with sparsentan in Phase 2 and 3 clinical trials^8, 9^, which demonstrated strong antiproteinuric effect across different phenotypes in patients with FSGS.

Urinary marker associations with the sparsentan score indicate the potential for non-invasively measuring intrarenal signaling activity of a putatively sparsentan-responsive network. The top correlated urine aptamers included C7, which has been reported to be selectively expressed in mesangial cells, NOTCH3 is important for maintain vascular function in the kidney^39–41^ and deletion in model systems reduces HIV-1 mediated renal injury, FGB (fibrinogen beta) is a component of fibrinogen, which is associated with CKD progression^42^, and UNC5B, a receptor for Netrin-1 expressed in endothelial cells that plays a role in regulating angiogenesis^43, 44^. The ability to measure different aspects of kidney health and CKD progression non-invasively may be key in identifying early responders to various therapies in clinical trials.

## Study Limitations

NEPTUNE is an observational study which we have used for cross-species analysis and as such, we cannot readily determine the underlying causality of the expression profiles in patients with FSGS with respect to AT_1_/ET-1 receptor activation. An interventional study with biopsies at enrollment and at study completion is beyond the scope of current work.

## Data Availability

All data produced in the present study are available upon reasonable request to the authors

## Acknowledgements

We thank Lalita Subramanian, PhD, for help with writing, editing, and formatting this manuscript. The Nephrotic Syndrome Study Network Consortium (NEPTUNE), U54-DK- 083912, is a part of the National Institutes of Health (NIH) Rare Disease Clinical Research Network (RDCRN), supported through collaboration between the Office of Rare Diseases Research, National Center for Advancing Translational Sciences, and the National Institute of Diabetes, Digestive, and Kidney Diseases (NIDDK). Additional funding and/or programmatic support for this project has also been provided by the University of Michigan, the NephCure Kidney International and the Halpin Foundation, and the Applied Systems Biology Core at the University of Michigan George M. O’Brien Kidney Translational Core Center (2P30-DK- 08194). LHM is supported through funding from NIH/NIDDK, K08 DK115891-01. NEPTUNE contributing members are listed in the Supplementary Acknowledgment.

## List of supplementary materials Supplementary acknowledgment

Supplementary Figure 1. Differential expression volcano plots and pathway enrichment of differentially expressed genes.

Supplementary Figure 2. Scatterplots to assess reversal of global transcriptional profiles in response to sparsentan in rat kidneys. Log2 transformed gene expression fold changes Sparsentan (ADR-Spa) treated compared to ADR-Veh kidneys (y-axis) were plotted against gene fold changes in ADR-Veh compared Veh kidneys (x-axis)

Supplementary Table 1. Rat-human ortholog mapping to identity genes used to generate the sparsentan activity score in transcriptomes from kidneys of patients with FSGS in the NEPTUNE cohort.

