## Supplementary text and supplementary figures for "Framework for precision medicine in focal segmental glomerulosclerosis: Translation of sparsentan-responsive genes in a rat model to kidney disease associated proteins in biofluids"

**List of supplementary materials**

**Supplementary acknowledgment**

**Supplementary Figure S1.** Differential expression volcano plots and pathway enrichment of differentially expressed genes.

**Supplementary Figure S2.** Scatterplots to assess reversal of global transcriptional profiles in response to sparsentan in rat kidneys. Log2 transformed gene expression fold changes Sparsentan (ADR-Spa) treated compared to ADR-Veh kidneys (y-axis) were plotted against gene fold changes in ADR-Veh compared Veh kidneys (x-axis)

**Supplementary Table S1.** Rat-human ortholog mapping to identity genes used to generate the sparsentan activity score in transcriptomes from kidneys of patients with FSGS in the NEPTUNE cohort.

### Supplementary acknowledgement

**Members of the Nephrotic Syndrome Study Network (NEPTUNE)**

**NEPTUNE Collaborating Sites**
*Atrium Health Levine Children's Hospital, Charlotte, SC*: Susan Massengill[*], Layla Lo[#]
*Cleveland Clinic, Cleveland, OH*: Katherine Dell[*], John O'Toole[*], John Sedor[**], Victoria Grange[#]
*Children's Hospital, Los Angeles, CA*: Ian Macumber[*], Alyssa Parry[#]
*Children's Mercy Hospital, Kansas City, MO*: Tarak Srivastava[*], Kelsey Markus[#]
*Cohen Children's Hospital, New Hyde Park, NY*: Christine Sethna[*], Suzanne Vento[#]
*Columbia University, New York, NY:* Pietro Canetta[*]
*Duke University Medical Center, Durham, NC:* Opeyemi Olabisi[*], Rasheed Gbadegesin[**], Maurice Smith[#]
*Emory University, Atlanta, GA:* Laurence Greenbaum[*], Chia-shi Wang[*], Emily Yun[#]
*The Lundquist Institute, Torrance, CA:* Sharon Adler[*], Janine LaPage[#]
*John H Stroger Cook County Hospital, Chicago, IL:* Amatur Amarah[*]
*Johns Hopkins Medicine, Baltimore, MD:* Meredith Atkinson[*], Sara Boynton[#]
*Mayo Clinic, Rochester, MN:* John Lieske, Marie Hogan, Fernando Fervenza
*Medical University of South Carolina, Charleston, SC:* David Selewski[*], Cheryl Alston[#]
*Montefiore Medical Center, Bronx, NY:* Kim Reidy[*], Michael Ross[*], Frederick Kaskel[**], Patricia Flynn[#]
*New York University Medical Center, New York, NY:* Laura Malaga-Dieguez[*], Olga Zhdanova[**], Laura Jane Pehrson[#], Melanie Miranda[#]
*The Ohio State University College of Medicine, Columbus, OH*: Salem Almaani[*], Laci Roberts[#]
*Stanford University, Stanford, CA:* Richard Lafayette[*], Shiktij Dave[#]
*Temple University, Philadelphia, PA:* Iris Lee[**]
*Texas Children's Hospital at Baylor College of Medicine, Houston, TX*: Shweta Shah[*], Sadaf Batla[# #]
*University Health Network Toronto:* Heather Reich[*], Michelle Hladunewich[**], Paul Ling[#], Martin Romano[#]
*University of California at San Francisco, San Francisco, CA*: Paul Brakeman[*], Daniel Schrader
*University of Colorado Anschutz Medical Campus, Aurora, CO*: James Dylewski[*] Nathan Rogers[#]
*University of Kansas Medical Center, Kansas City, KS*: Ellen McCarthy[*], Catherine Creed[#]
*University of Miami, Miami, FL:* Alessia Fornoni[*], Miguel Bandes[#]
*University of Michigan, Ann Arbor, MI:* Matthias Kretzler[*], Laura Mariani[*], Zubin Modi[*], A Williams[#], Roxy Ni[#]
*University of Minnesota, Minneapolis, MN:* Patrick Nachman[*], Michelle Rheault[*], Amy Hanson[#], Nicolas Rauwolf[#]
*University of North Carolina, Chapel Hill, NC:* Vimal Derebail[*], Keisha Gibson[*], Anne Froment[#], Mary Mac McGown Collie[#]
*University of Pennsylvania, Philadelphia, PA:* Lawrence Holzman[*], Kevin Meyers[**], Krishna Kallem[#], Aliya Edwards[#]
*University of Texas San Antonio, San Antonio, TX*: Samin Sharma[**]
*University of Texas Southwestern, Dallas, TX:* Elizabeth Roehm[*], Kamalanathan Sambandam[**], Elizabeth Brown[**], Jamie Hellewege
*University of Washington, Seattle, WA:* Ashley Jefferson[*], Sangeeta Hingorani[**], Katherine Tuttle[**§], Linda Manahan [#], Emily Pao[#], Kelli Kuykendall[§]
*Wake Forest University Baptist Health, Winston-Salem, NC:* Jen Jar Lin[**]
*Washington University in St. Louis, St. Louis, MO*: Vikas Dharnidharka[*]

**Data Analysis and Coordinating Center:** *University of Michigan:* Matthias Kretzler[*], Brenda Gillespie[**], Laura Mariani[**], Zubin Modi[**], Eloise Salmon[**], Howard Trachtman[**], Tina Mainieri, Gabrielle Alter, Michael Arbit, Hailey Desmond, Sean Eddy, Damian Fermin, Wenjun Ju, Maria Larkina, Chrysta Lienczewski, Rebecca Scherr, Jonathan Troost, Amanda Williams, Yan Zhai; *Arbor Collaborative for Health:* Colleen Kincaid, Shengqian Li, Shannon Li; *Cleveland Clinic:* Crystal Gadegbeku[**], *Duke University:* Laura Barisoni[**]; John Sedor[**], *Harvard University:* Matthew G Sampson[**]; *Northwestern University:* Abigail Smith[**]; *University of Pennsylvania:* Lawrence Holzman[**], Jarcy Zee[**]

**Digital Pathology Committee:** Carmen Avila-Casado *(University Health Network)*, Serena Bagnasco *(Johns Hopkins University)*, Lihong Bu *(Mayo Clinic)*, Shelley Caltharp *(Emory University)*, Clarissa Cassol *(Arkana)*, Dawit Demeke *(University of Michigan)*, Brenda Gillespie *(University of Michigan)*, Jared Hassler *(Temple University)*, Leal Herlitz *(Cleveland Clinic)*, Stephen Hewitt *(National Cancer Institute)*, Jeff Hodgin *(University of Michigan)*, Danni Holanda *(Arkana)*, Neeraja Kambham *(Stanford University)*, Kevin Lemley, Laura Mariani *(University of Michigan)*, Nidia Messias *(Washington University)*, Alexei Mikhailov *(Wake Forest)*, Vanessa Moreno *(University of North Carolina)*, Behzad Najafian *(University of Washington)*, Matthew Palmer *(University of Pennsylvania)*, Avi Rosenberg *(Johns Hopkins University)*, Virginie Royal *(University of Montreal)*, Miroslav Sekulik *(Columbia University)*, Barry Stokes *(Columbia University)*, David Thomas *(Duke University)*, Ming Wu *(University of New York)*, Michifumi Yamashita *(Cedar Sinai)*, Hong Yin *(Emory University)*, Jarcy Zee *(University of Pennsylvania)*, Yiqin Zuo *(University of Miami)*. Co-Chairs: Laura Barisoni *(Duke University)*, Cynthia Nast *(Cedar Sinai)*.

Supplemental Figure 1

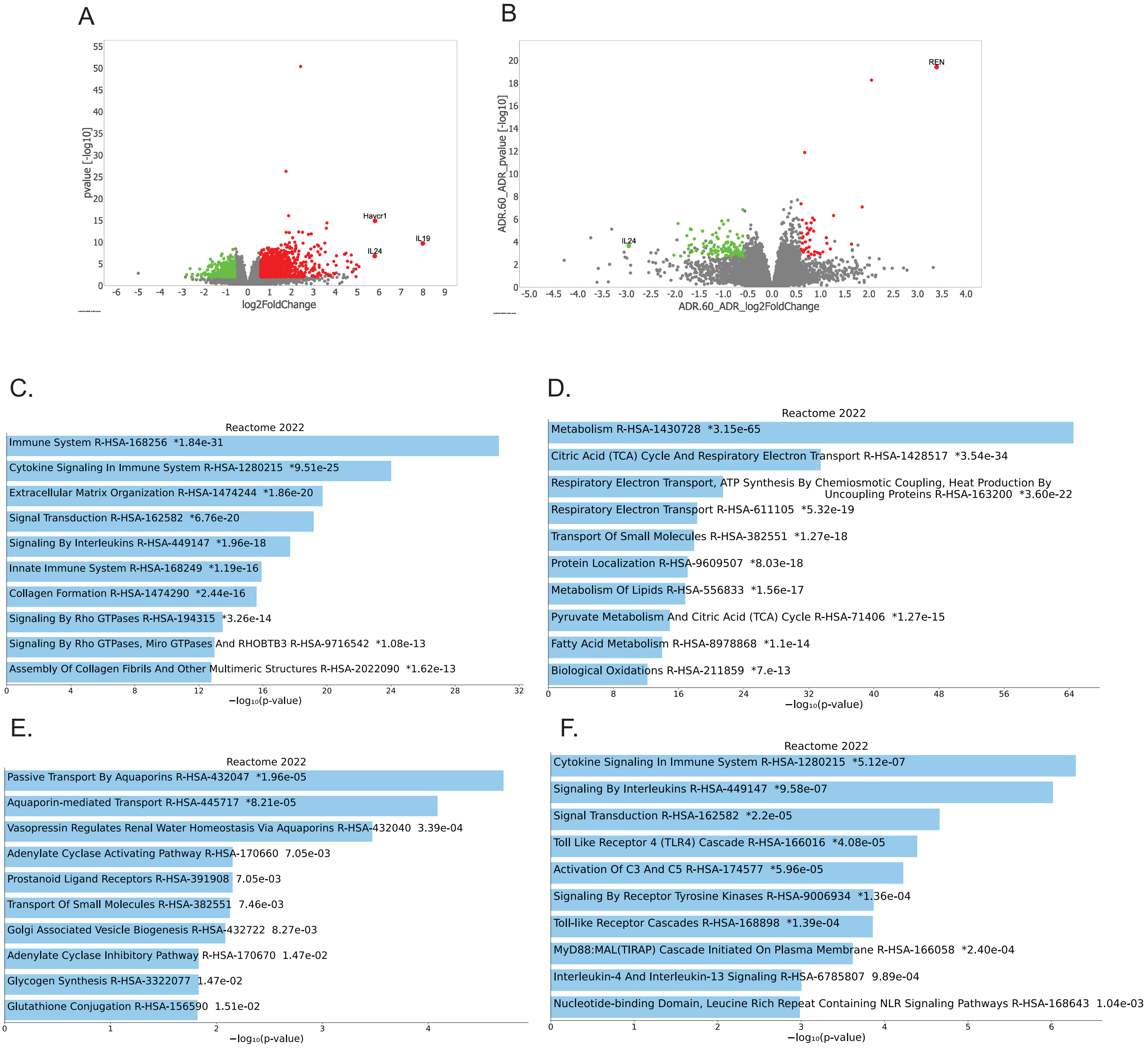


**Supplementary Figure S1.Volcano plots of differentially expressed genes included for pathway analysis** (red=up-regulated, green=down-regulated) for the A) ADR-Veh vs. Veh comparison to establish the disease in the model and B) ADR-Spa60 vs. ADR-Veh comparison to establish the effects of sparsentan in the context of disease in the model. C) Enrichment analysis results of genes up-regulated and D) genes down-regulated from A 60 (ADR-Veh vs. Veh) and E) Enrichment analysis results of genes up-regulated and F) genes down-regulated in B (ADR-Spa60 vs. ADR-Veh).

Supplementary Figures S2

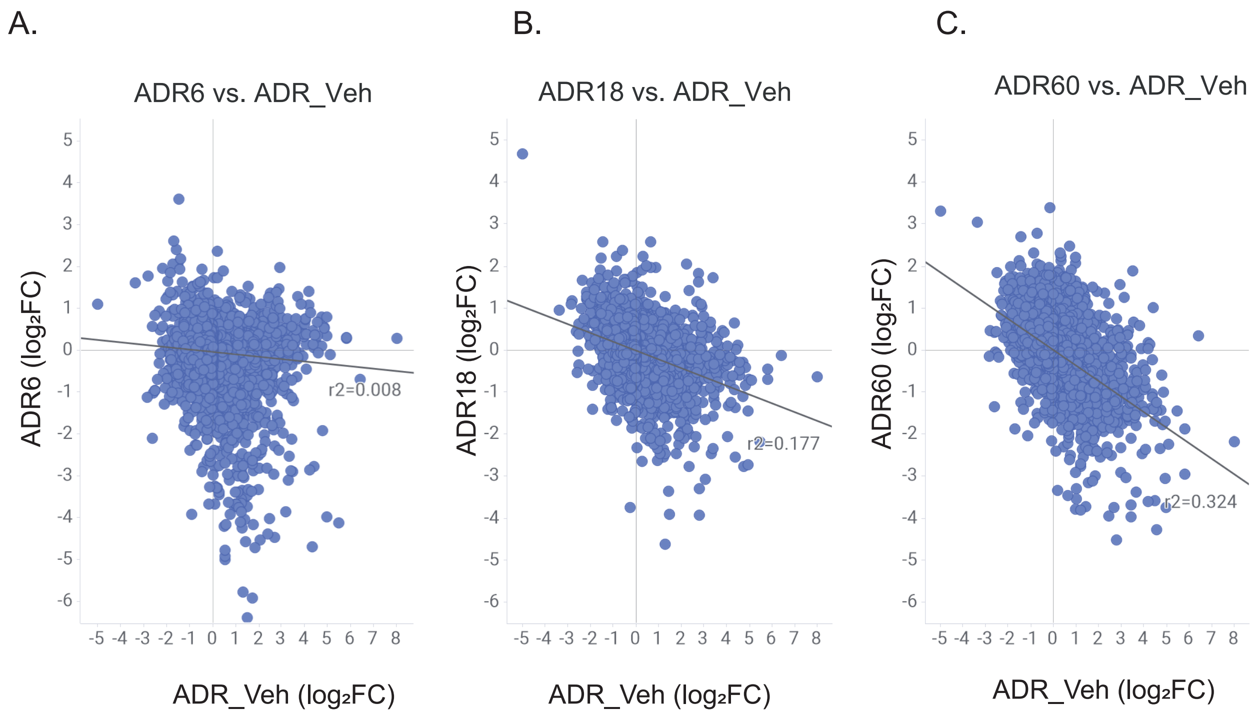


**Supplementary Figure S2. Comparison of the totality of gene expression log2 fold changes in each sparsentan treated animal group in comparison to the model.** Each spot represents an individual gene A) ADR-Spa6 vs. ADR-Veh log2 FC compared to ADR-Veh vs. Veh log2 FC (r=-0.09), B) ADR-Spa18 vs. ADR-Veh log2 FC compared to ADR-Veh vs. Veh log2 FC (r=-0.42), and C) ADR-Spa60 vs. ADR-Veh log2 FC vs. ADR-Veh vs. Veh log2 FC (r=-0.57).
